# Multimodal surface coils for low field MR imaging

**DOI:** 10.1101/2024.04.14.24305802

**Authors:** Yunkun Zhao, Aditya A Bhosale, Xiaoliang Zhang

## Abstract

Low field MRI is safer and more cost effective than the high field MRI. One of the inherent problems of low field MRI is its low signal-to-noise ratio or sensitivity. In this work, we introduce a multimodal surface coil technique for signal excitation and reception to improve the RF magnetic field (B_1_) efficiency and potentially improve MR sensitivity. The proposed multimodal surface coil consists of multiple identical resonators that are electromagnetically coupled to form a multimodal resonator. The field distribution of its lowest frequency mode is suitable for MR imaging applications. The prototype multimodal surface coils are built, and the performance is investigated and validated through numerical simulation, standard RF measurements and tests, and comparison with the conventional surface coil at low fields. Our results show that the B_1_ efficiency of the multimodal surface coil outperforms that of the conventional surface coil which is known to offer the highest B_1_ efficiency among all coil categories, i.e., volume coil, half-volume coil and surface coil. In addition, in low-field MRI, the required low-frequency coils often use large value capacitance to achieve the low resonant frequency which makes frequency tuning difficult. The proposed multimodal surface coil can be conveniently tuned to the required low frequency for low-field MRI with significantly reduced capacitance value, demonstrating excellent low-frequency operation capability over the conventional surface coil.

## 1. Introduction

Magnetic resonance imaging (MRI) is a promising tool to provide detailed images depicting the internal structures [1-6], functions [7-14] and metabolic processes [15-23] of the living system without the use of ionizing radiation. Traditional high-field MRI systems, typically operating at 1.5 Tesla or higher, are prevalent in clinical settings due to their high signal-to-noise ratio (SNR), which contributes to their ability to produce high-resolution images and increased spectral dispersion [24-37]. However, MRI systems operating at higher field strengths are often associated with high operating costs [38, 39], substantial power requirements [40-44], radio frequency (RF) challenges [45-61] and potential safety concerns, especially for patients with certain medical implants or conditions that contraindicate exposure to strong magnetic fields [62, 63]. In recent years, there has been a growing interest in low-field MRI systems, defined as those operating at magnetic field strengths in the range from 0.25 Tesla to 1.0 Tesla [64-66]. These systems offer several advantages over their high-field counterparts, including lower operating costs, reduced power consumption, and improved safety profile, making them more accessible and suitable for a wider range of patients [64-68].

A major challenge associated with low field MRI is the inherently low signal-to-noise ratio (SNR) [69-72], which significantly limits the spatial/temporal resolution and specificity of the image [73-75]. It is known that the MR SNR is directly related to the RF magnetic field B_1_, showing a linear relationship [69]. Therefore, improving the efficiency of the B_1_ field generated by RF coils may be a possible way to improve the SNR.

Conventional surface coils, which provide the highest B_1_ efficiency among all RF coil types (i.e. surface coil, volume coil and half-volume coil), still show limitations in providing sufficient B_1_ fields for signal reception/excitation in many imaging applications, resulting in suboptimal image sensitivity and resolution. To address this challenge, in this work we introduce and investigate an innovative solution: the multimodal surface coil. This novel design pushes the B_1_ boundary and significantly improves the B_1_ fields over those provided by conventional surface coils, with the potential to significantly enhance MR SNR. The core concept of the multimodal surface coil is based on a set of stacked resonators that are electromagnetically coupled to form a multimodal RF resonator [76]. This design not only improves B_1_ efficiency, but also incorporates a low frequency tuning capability. Such a feature is particularly advantageous in low-field MRI, where tuning at the corresponding low frequencies poses a notable challenge. To validate our design, we have performed extensive full-wave electromagnetic simulations alongside standard RF bench tests and measurements. The proposed design is further validated by a comparison study with a conventional surface coil.

## 2. Methods

### 2.1 EM simulation

Figure 1 displays the simulation model of a multimodal surface coil. This coil is comprised of seven resonators or coil loops constructed with 6.35 mm wide copper tape. Six of these are identical square coils, each with a side length of 10 cm and equipped with a 60pF capacitance tuning capacitor for tuning to 42 MHz. The central coil contains an impedance matching circuit essential for driving the multimodal surface coil. A spacing of 5 mm between the coils is designed to enhance mutual inductive coupling, with the entire stacked assembly reaching a height of 3 cm. Our design, featuring multiple coils, inherently supports four resonant modes within the coupled stack. We have chosen to utilize the lowest resonant mode for imaging applications due to its superior field strength efficiency, a critical factor in optimizing image quality in MRI. A conventional surface coil has been used as a comparative setup. Mirroring the multimodal resonator, it was also defined by a 10 cm length. All designs, including the multimodal surface coil, were built using a 6.35 mm wide copper sheet conductor, tuned to 21.3 MHz, and impedance-matched to 50 ohms. In comparison study, the proposed multimodal surface coil ad conventional surface coil are placed 1 cm below a tank phantom with a dimension of 20×10×20 cm^2^. The conductivityσ= 0.39 S/m and the relative permittivity value ε_r_ = 188.76 are used in the simulation phantom to imitate the human brain tissue properties at 21 MHz. Performance assessments of the multimodal surface coil involved analyzing scattering parameters, and B_1_ efficiency, using field distribution plots. All electromagnetic field plots were normalized to 1 W of accepted power. Numerical results of the proposed designs were obtained using the electromagnetic simulation software CST Studio Suite (Dassault Systèmes, Paris, France).

**Figure 1.**
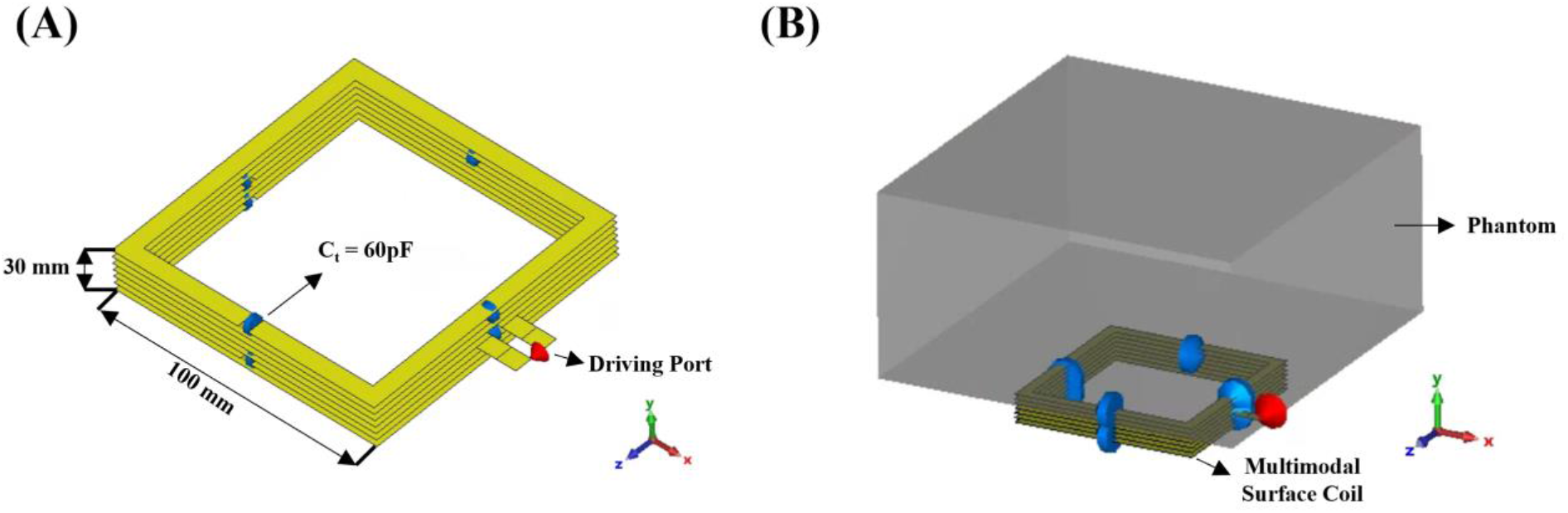
(A) Simulation model and size of multimodal surface coil. (B) Proposed multimodal surface coil loaded with a cuboid phantom.

### 2.2 Bench Test Model Assembly

Figure 2 shows photographs and dimensions of bench test models of the multimodal surface coil and conventional surface coils. The bench test models have the same dimensions as the simulation model. The conductors of the multimodal surface coils were built with 6.35 mm wide copper tape and on a 3D printed polylactide structure. The imaging resonant frequency was tuned to 21 MHz and matched to 50 ohms by careful selection of the capacitance value on each coil. We used 7 identical fixed-tuning capacitors with 39 pF capacitance. The matching circuit was implemented as shown in Figure 2A. One capacitor with 330 pF connected in parallel to the feeding line was employed for impedance matching.

**Figure 2.**
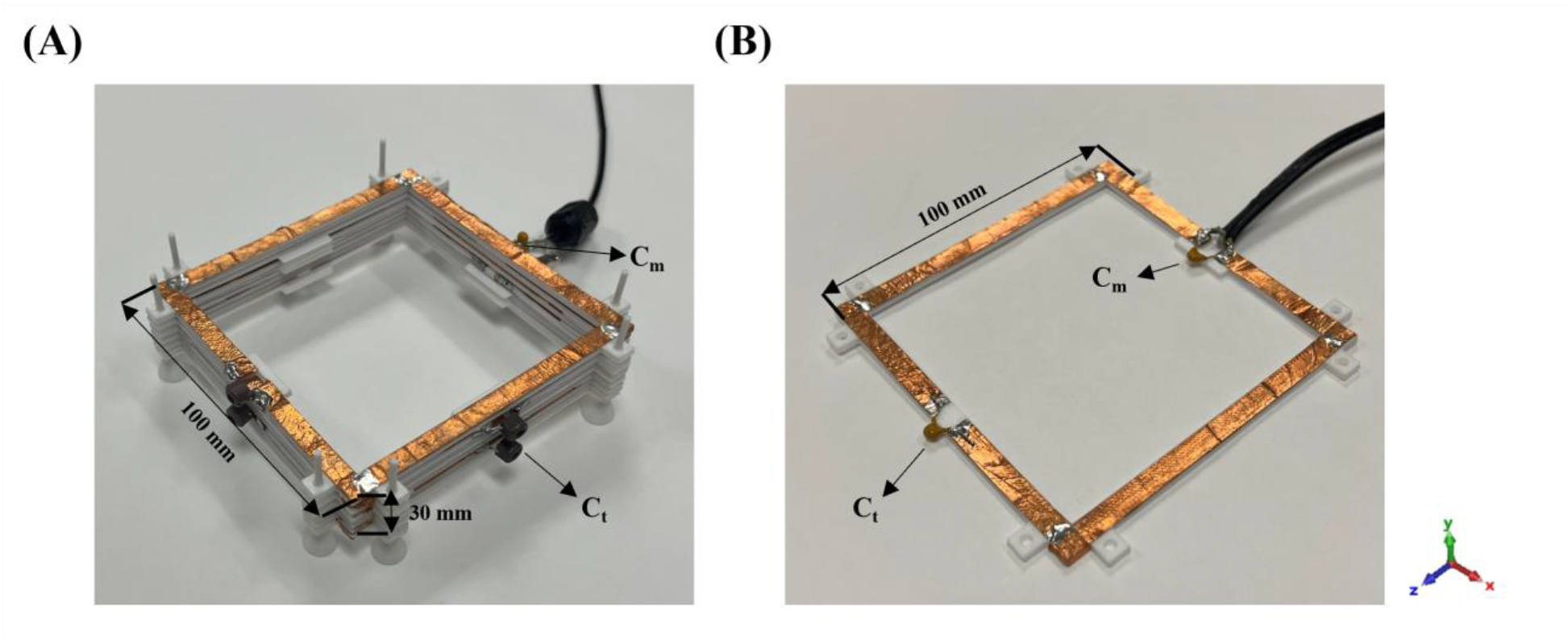
(A) Bench test model of multimodal surface coil. (B) Bench test model of conventional surface coil.

For comparison, a conventional surface coil [15] has also been made. The conventional surface coil has the same dimensions as their simulation model and the multimodal surface coil. It was also built using 6.35 mm width thick copper tape on a 3D printed polylactide structure and was tuned to 21 MHz and matched to 50 ohms by tuning capacitors and a matching circuit. The B_1_ field strength and distribution have been visualized by a sniffer positioning system combined with a magnetic and electric field measurement setup shown in Figure 3 in an unloaded case. The measurement setup includes a sniffer (H-field probe) which integrated to a high-resolution router machine (Genmitsu CNC PROVerXL 4030) for positioning of the field probe and detect the B_1_ field generated by the RF coil in 3D space. The probe is connected to a vector network analyzer (VNA) from Keysight, E5061B, Santa Clara, CA, USA. The raw data, including the output and accepted power and scattering parameters acquired from VNA, will be transmitted to computer and processed by MATLAB to calculate the B_1_ field efficiency map. The B_1_ field efficiency map is obtained for a 10 × 10 cm^2^ X-Z plane slice located 1 cm above the RF coils and a 7 × 10 cm^2^ X-Y plane slice located 2 cm above the RF coils with each measurement point spaced at intervals of 2.5 mm. All results were normalized to 1 W of accepted power.

**Figure 3.**
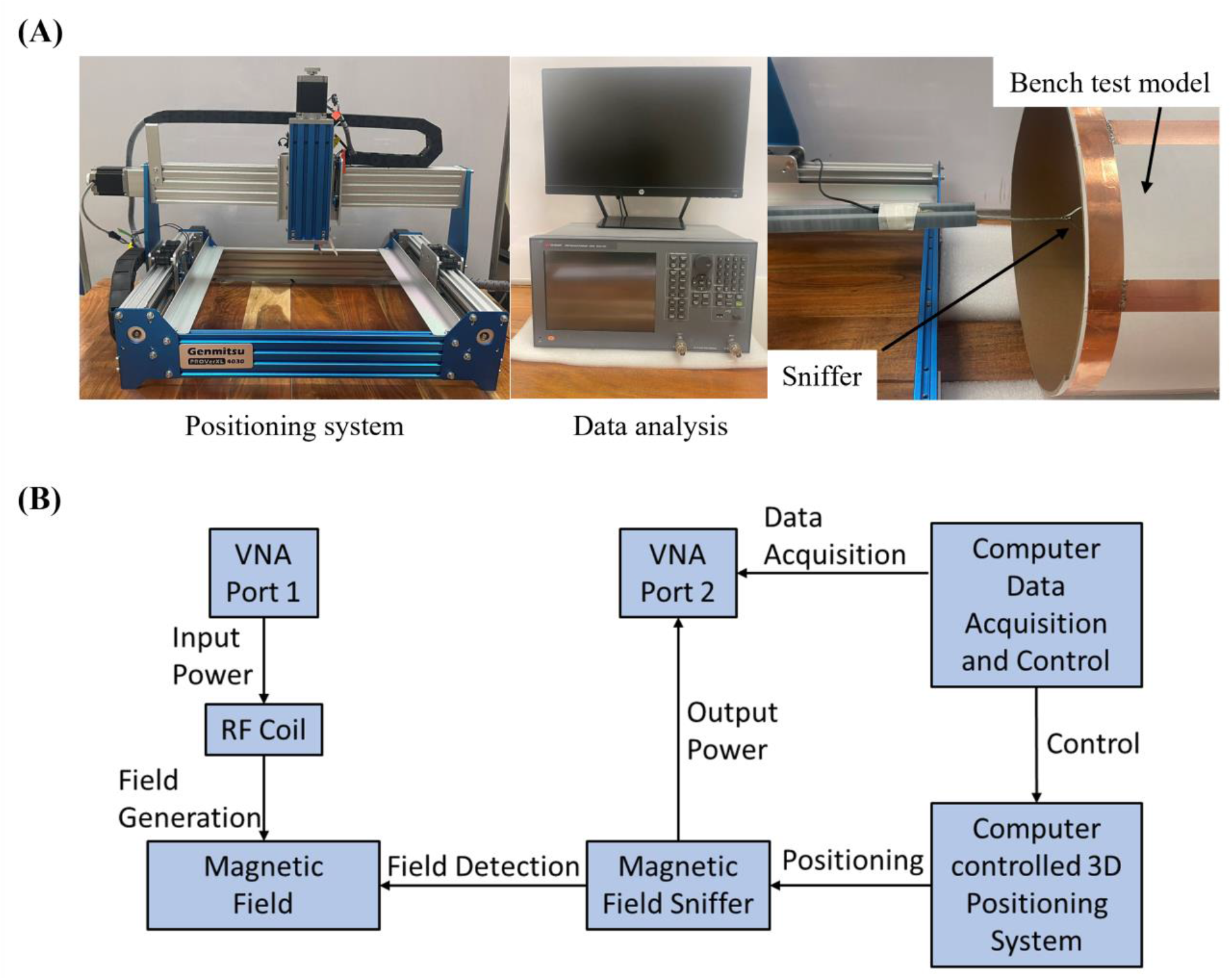
(A) Photograph of measurement setup including the sniffer-positioning system, network analyzer and the data process computer. (B) Schematic of the measurement workflow to obtain B_1_ field efficiency map.

## 3. Results

### 3.1 Simulated Resonant Frequency and Field Distribution

Figure 4 presents the simulated scattering parameters vs frequency for the multimodal surface coil. This graph indicates the emergence of strong coupling between the coils, which manifests as four distinct split resonant peaks. The highest of these peaks occurs at 79 MHz, while the lowest, which is pertinent for imaging purposes, is observed at 21 MHz. It is noteworthy that each coil in the originally resonates at 42 MHz, underscoring the effectiveness of our tuning approach using a relatively low capacitance value to achieve a lower resonant frequency for a coil of the same size. In comparison, a conventional surface coil of the same size requires a 200pF capacitance to reach 21 MHz, leading to less accurate and convenient frequency tuning. In Figure 5, we illustrate the B_1_ field efficiency maps across Y-Z, X-Z, and X-Y planes within the phantom, positioned at the center of the axis. These plots visualize the B_1_ field efficiency of the multimodal surface coils. The results from these simulations demonstrate that our multimodal surface coil exhibits a stronger B field efficiency while maintaining a B_1_ field distribution comparable to that of a conventional surface coil.

**Figure 4.**
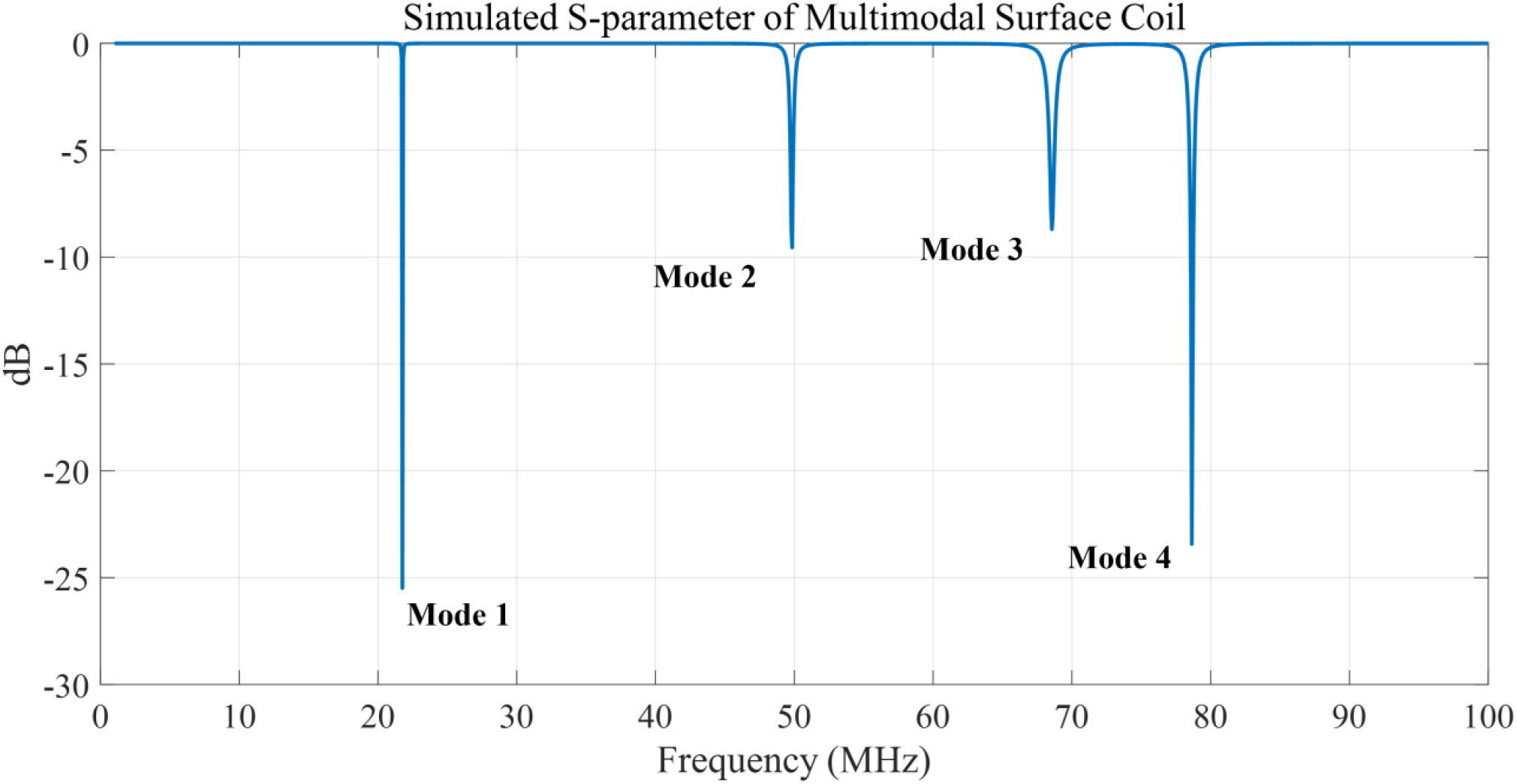
Simulated scattering parameters vs. frequency of the multimodal surface coil.

**Figure 5.**
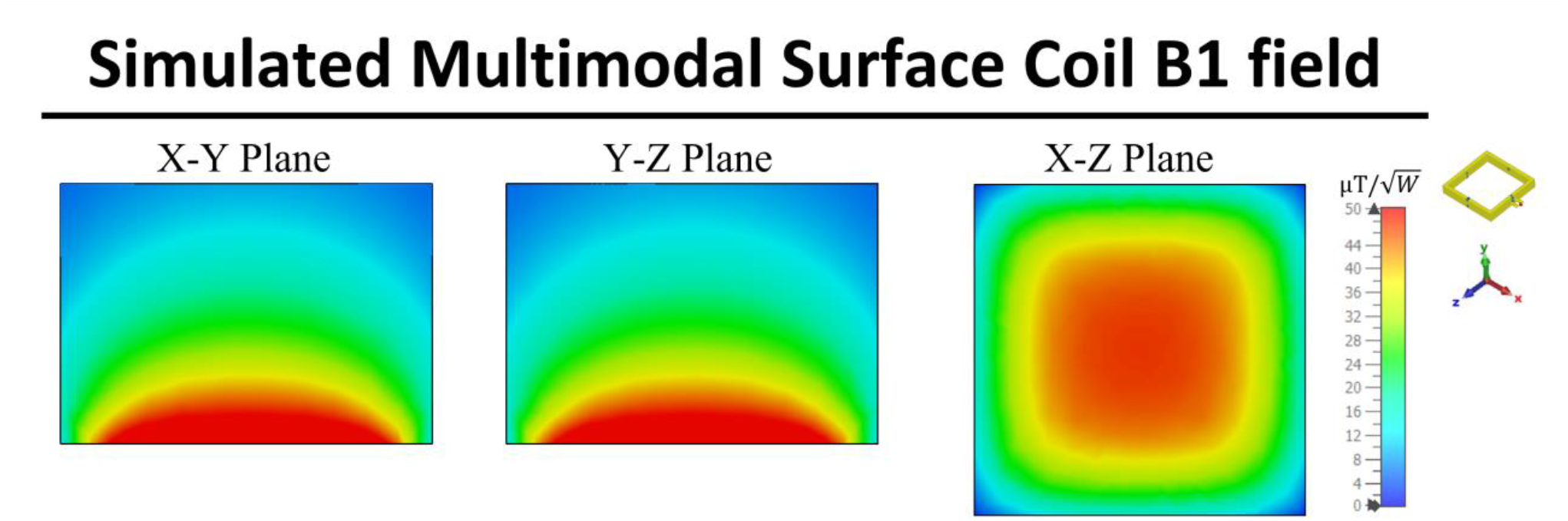
Simulated Y-Z, X-Y, and X-Z plane B field efficiency maps inside phantom generated by multimodal surface coils

### 3.2 Measured Scattering Parameters and Field Distribution

Figure 6 shows that the S-parameter vs. frequency plots of the coupled stack-up coil are in good agreement with the simulation results. Four resonant modes with 21.8 MHz, 58.9 MHz, 73.6 MHz, and 85.4 MHz were formed. Figure 7 shows the B_1_ field efficiency distribution map on X-Y, Y-Z, and X-Z planes measured with a 3-D magnetic field mapping system. Multimodal surface coil shows strong B field efficiency and similar field distribution pattern on all three planes and is in accordance with the simulation result, which also indicates that the simulation results are accurate and reliable.

**Figure 6.**
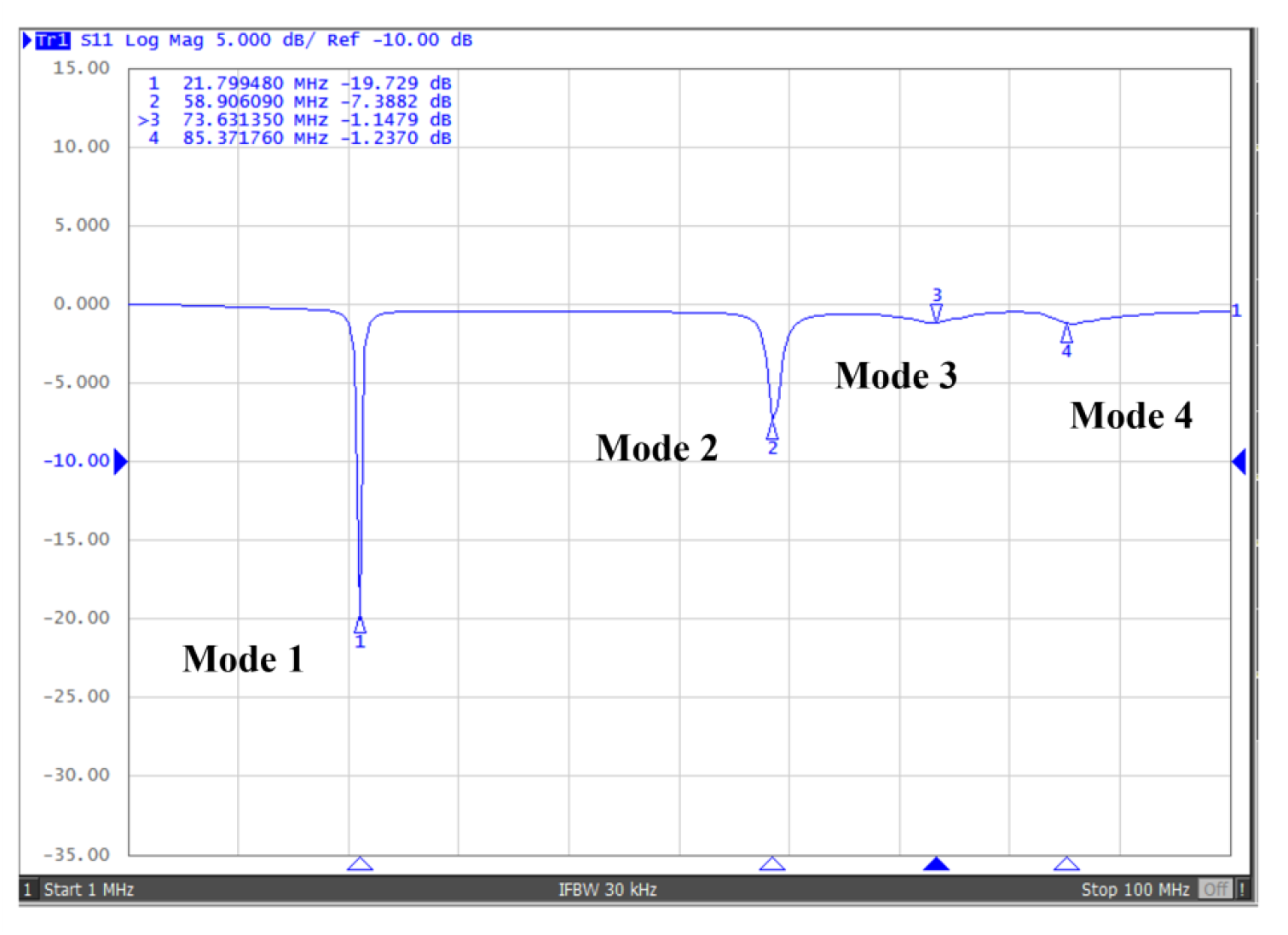
Scattering parameters vs. frequency of the bench test model of multimodal surface coils.

**Figure 7.**
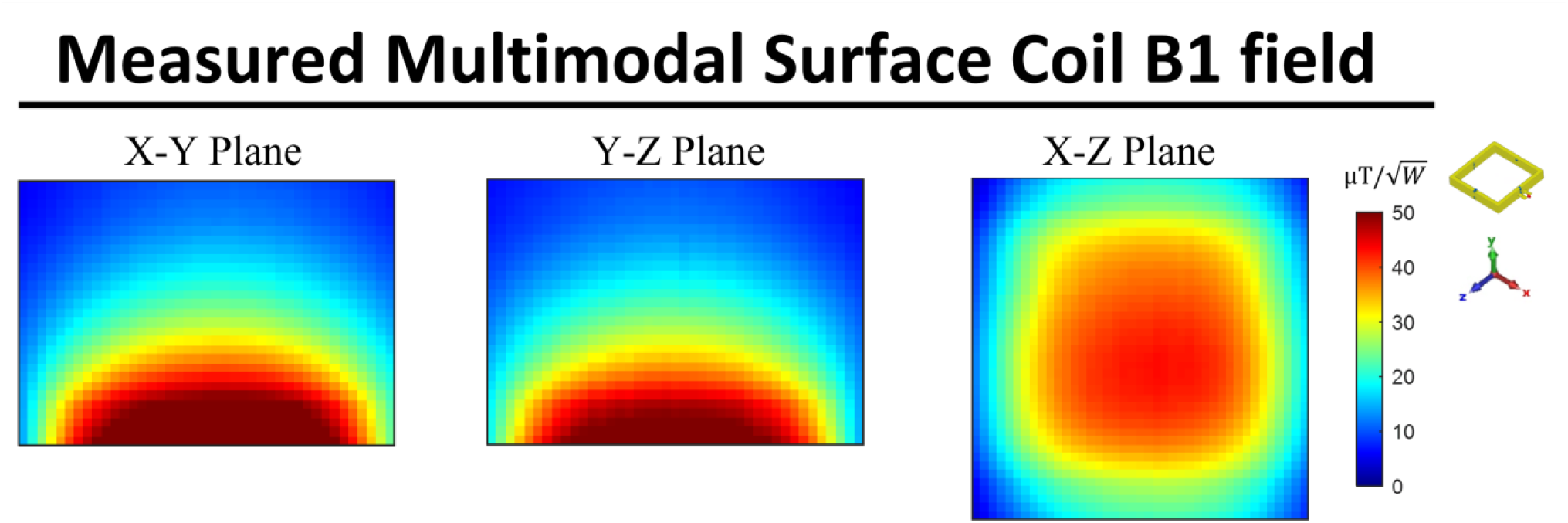
Measured B field efficiency maps on the X-Y, Y-Z, and X-Z plane of multimodal surface coil.

### 3.3 Field Distribution and Efficiency Evaluation

Figure 8 provides a comparative analysis of the simulated B_1_ field efficiency between the multimodal surface coil, and a conventional surface coil. The results demonstrate that, despite maintaining a field distribution similar to that of the conventional surface coil, the multimodal surface coil exhibits a significantly higher B_1_ field efficiency. This finding is crucial as it underscores the effectiveness of the multimodal design in improving the quality of MRI imaging. In Figure 7B, we present a 1-D plot of the B_1_ field efficiency, correlating to the vertical and horizontal dashed lines depicted in Figure 7A. This graphical representation provides a detailed insight into the spatial efficiency of the B_1_ field. Notably, the average B_1_ field efficiency generated by the multimodal surface coil surpasses that of the conventional surface coil by 46.8% along the X-axis, measured at a position 2 cm above the coil.

**Figure 8.**
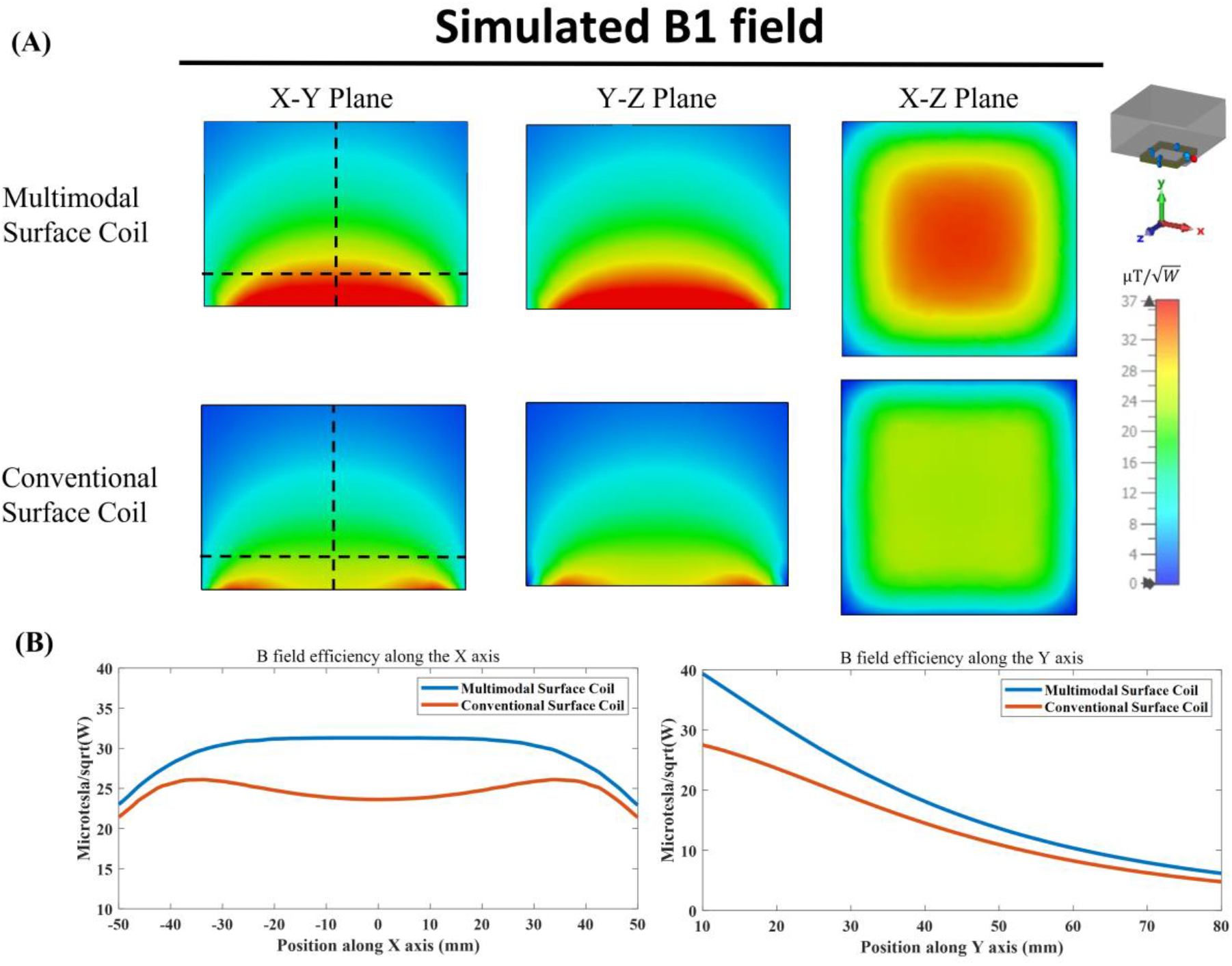
(A) Simulated Y-Z, X-Y, and X-Z plane B field efficiency maps inside phantom generated by multimodal surface coils, and conventional surface coil. Y-Z and X-Y planes are at the center of the coil and X-Z plane is 2 cm above the coil. (B) 1-D plot of B_1_ field efficiency along the vertical and horizontal dashed line shown in (A).

Figure 9 presents the comparison of the B field efficiency between the bench test model of the multimodal surface coil and the conventional coil. The measured B-field efficiency distribution and strength are found to be in alignment with our simulation results. This congruence is vital as it validates our design, confirming that the multimodal surface coil possesses a stronger B_1_ field efficiency within the imaging area compared to the conventional surface coil.

**Figure 9.**
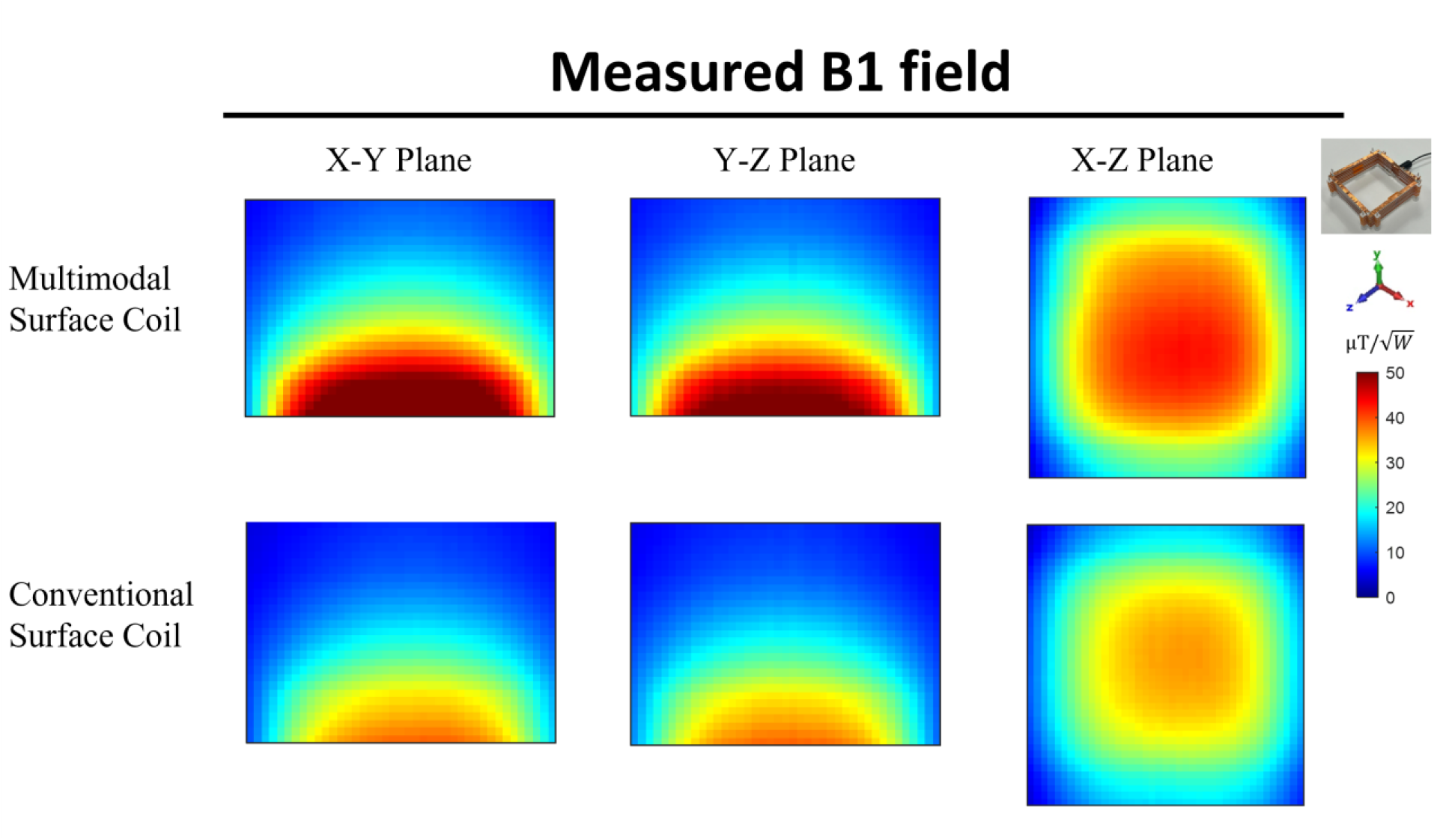
Comparison between measured B field efficiency maps on the X-Y, Y-Z, and X-Z plane of multimodal surface coils and conventional surface coil.

## 4. Discussion

Due to the use of the lowest frequency mode that possesses an MR-demanded field distribution, the multimodal surface coil demonstrates the excellent capability for low frequency or low field MR imaging applications in humans. To increase the resonant frequency of the proposed multimodal surface coil to exploit its potential in high field MR imaging applications, one tangible way that can be applied is to reduce the inductance of the circuit. This can be achieved by reducing the size of the coil. With the increased frequency by downsizing the coil, it is possible to generate a more efficient reception/excitation approach using the proposed multimodal surface coil technique for high field small animal MR imaging with expected higher sensitivity over the traditional surface coils [77].

If an appropriate decoupling technique is applied, the proposed multimodal surface coils can potentially be used as building blocks to design multichannel coil arrays to either improve SNR and imaging coverage, or to enable parallel imaging for accelerated signal excitation and reception. Due to their multimodal or multi-resonance nature, decoupling of multimodal coil elements could be a challenging task. The magnetic wall or ICE decoupling technique could be a working method for this electromagnetic decoupling need due to its unique broadband decoupling capability. In addition, high impedance design can also potentially be used to achieve the required inter-coil decoupling in multichannel coil arrays.

## 5. Conclusion

In this work, a multimodal surface coil design has been proposed and the prototype multimodal surface coil has been successfully developed for low field MR imaging applications. The proposed multimodal surface RF coil has demonstrated significant improvements in B_1_ efficiency over conventional surface coils, potentially leading to improved SNR for low field MRI, where low SNR is a major drawback, limiting spatial and temporal resolutions and spectral dispersion. Due to the use of the lowest frequency mode, the proposed multimodal surface coil design also helps to achieve low frequency tuning which is technically challenging at low magnetic fields.

## Data Availability

All data produced in the present work are contained in the manuscript

## Acknowledgments

This work is supported in part by the NIH under a BRP grant U01 EB023829 and by the State University of New York (SUNY) under SUNY Empire Innovation Professorship Award.

